# Detection of SARs-CoV-2 in wastewater, using the existing environmental surveillance network: An epidemiological gateway to an early warning for COVID-19 in communities

**DOI:** 10.1101/2020.06.03.20121426

**Authors:** Salmaan Sharif, Aamer Ikram, Adnan Khurshid, Muhammad Salman, Nayab Mehmood, Yasir Arshad, Jamal Ahmad, Rana Muhammad Safdar, Mehar Angez, Muhammad Masroor Alam, Lubna Rehman, Ghulam Mujtaba, Jaffar Hussain, Johar Ali, Ribqa Akthar, Muhammad Wasif Malik, Zeeshan Iqbal Baig, Muhammad Suleman Rana, Muhammad Usman, Muhammad Qaisar Ali, Abdul Ahad, Nazish Badar, Massab Umair, Sana Tamim, Asiya Ashraf, Faheem Tahir, Nida Ali

## Abstract

**Background:** The ongoing COVID-19 pandemic caused by SARs-CoV-2 was transmitted person to person via droplet infections and fecal-oral transmission. This illustrates the probability of environmentally facilitated transmission, mainly the sewage.

**Method:** We used existing Pakistan polio environment surveillance network to investigate presence of SARs-CoV-2 using three commercially available kits and E-Gene detection published assay for surety and confirmatory of positivity. A Two-phase separation method is used for sample clarification and concentration. An additional high-speed centrifugation (14000Xg for 30 min) step was introduced, prior RNA extraction, to increase viral RNA yield resulting a decrease in *Cq* value.

**Results:** A total of 78 wastewater samples collected from 38 districts across Pakistan, 74 wastewater samples from existing polio environment surveillance sites, 3 from drains of COVID-19 infected areas and 1 from COVID 19 quarantine center drainage, were tested for presence of SARs-CoV-2. 21 wastewater samples (27%) from 13 districts turned to be positive on RT-qPCR. SARs-COV-2 RNA positive samples from areas with COVID patients and COVID 19 patient quarantine center drainage strengthen the findings and use of wastewater surveillance in future. Furthermore, sequence data of partial ORF 1a generated from COVID 19 patient quarantine center drainage sample also reinforce our findings that SARs-CoV-2 can be detected in wastewater.

**Discussion:** This study finding indicates that SARs-CoV-2 detection through wastewater surveillance has an epidemiologic potential that can be used as early warning system to monitor viral tracking and circulation in cities with lower COVID-19 disease burden or heavily populated areas where door-to-door tracing may not be possible. However, attention needed on virus concentration and detection assay to increase the sensitivity. Development of highly sensitive assay will be an indicator for virus monitoring and to provide early warning signs.

## Introduction

Novel coronavirus pneumonia (COVID-19) caused by SARS-Cov-2 infection has become a global emergency through its widespread infection with 6,189,560 confirmed cases resulting 372,469 deaths in 213 countries as of 1^st^ June, 2020 (JHU 2020). In December 2019, cluster of pneumonia like disease cases with symptoms including fever, difficulty in breathing, cough and invasive lesion on both lungs were reported from Wuhan, China (WHO 2020a). The causative agent was identified as a Severe acute respiratory syndrome coronavirus 2 (SARS-CoV-2) after ruling out SARS-CoV, MERS-CoV, influenza, avian influenza, adenovirus and other common respiratory pathogens (WHO 2020b). Coronaviruses belonging to family *Cornaviridae* are enveloped, non-segmented positive sense RNA viruses distributed in human and mammals (Richman DD 2016). However, majority of human coronaviruses have mild infections but two betacornaviruses; severe acute respiratory syndrome coronavirus (SARS-CoV) and Middle East respiratory syndrome coronavirus (MERS-CoV) caused outbreaks in last two decades with 10% and 35.6% mortality rate respectively (de Groot et al. 2013; Hui et al. 2004; Ramadan and Shaib 2019).

. On March 11, 2020 WHO declared it as pandemic, when disease was reported in 114 countries (WHO 2020c). The primary routes of viral transmission (SARS-CoV-2) was considered to be through droplet infections and person to person close contact, but later it is evident from various published studies that there is increasing possibility of fecal-oral transmission (Adhikari et al. 2020; Cheng et al. 2004). This shows the probability of environmentally mediated transmission. Since the early days of pandemic, we got interested in understanding and utilizing the role of environmental sampling mainly the sewage.

SARS-CoV-2 resembles 82% with SARS coronavirus (SARS-CoV) which caused an outbreak in 2003. Studies have shown the survival of SARS-CoV in stool for up to 4 days (Hui et al. 2004). Another study described the presence of SARS-CoV and its infectious nature in water and sewage for days to weeks (Casanova et al. 2009). It was also described that faulty sewage system contaminated with SARs-CoV in a high-rise housing estate of Hong Kong during 2003 was linked to SARS outbreak involving large number of residents of surrounding buildings (Peiris et al. 2003). A recent study highlighted the shedding through stool of SARS-CoV-2 in cluster of 9 nCOVID-19 patients. It was reported that that the RNA concentration decreased from 10 RNA copies/g to 10 RNA copies/g after one week of symptom onset to third week (Roman Woelfel 2020). Since the source of transmission of SARS-CoV-2 is still unknown therefore wastewater transmission pathway can become an important mode (UNICEF 2020). Hence, the presence of SARS-CoV-2 in contaminated sewage sample and its role in transmission needs to be investigated. In this study, we used the existing polio environment surveillance network in Pakistan through which sewage samples were collected from designated sites in different districts of the country to investigate presence of poliovirus, its spread and molecular epidemiology. Same samples were processed and tested for detection of SARs-CoV-2 RNA.

## Methods

Untreated wastewater samples (sewage samples) selected for testing in this study were collected using the grab sampling technique. Most of them were those collected for routine polio environment surveillance (ES). Polio ES sites are either open drains or pumping stations and are sampled routinely on monthly basis. Each sampling site represent 100,000 – 300,000 population (WHO 2015). Besides, wastewater from drains of some areas with recent history of SARS-CoV2 cases were also collected for detection and re-confirmation of SARs-CoV-2 detection. Sampling personnel strictly followed the standard safety guidelines for personnel protective equipment (PPE) required for wastewater sampling. One liter of sewage water was collected from the mid-stream into a sterile, leak proof container at a downstream sampling site during the peak morning flow. These samples were transported in properly sealed container with information form, indicating sampling site, district, sampling date and sampling time, to laboratory within 48 hours of collection maintaining reverse cold chain (WHO 2015). Samples were processed in laboratory for virus concentration using the two-phase separation method (WHO 2003).

500 ml of each raw sewage specimens was concentrated. Firstly, clarification of the sample was done by pelleting of larger suspended solids by high speed centrifugation. The clarified sewage sample was mixed with defined amounts of polymers, dextran and polyethylene glycol (PEG). The homogenous mixture obtained by vigorous shaking is left to stand overnight at 4°C in a separation funnel. The polymer helped to form two distinct layers (phases) in the funnel which were collected and mixed with pellet formed in first step which was then treated with chloroform (WHO 2003) and further used for extraction of RNA.

Before proceeding directly for RNA extraction, an additional step was introduced to increase the yield of RNA, in contrast with direct processing. 400µl of processed sample was centrifuged at high speed (14000Xg) for 30 min to pellet the suspended solid particles. Virus may be partly bound to these solids. Supernatant was discarded carefully without disturbing the pellet, which was later used for RNA extraction.

Spin star viral nucleic acid kit 1.0 (ADT Biotech, Phileo Damansara 1, Petaling Jaya Part No. 811803) was used to extract the viral RNA. Internal control provided with kits were added as an amplification control in rRT-PCR. Pellet was dissolved with 430µl of lysis buffer supplied with kit, followed by 5 min vortexing to homogenously dissolve the pallet. Further processing was done as per the manufacturer’s instructions. The final elution volume is 60μl. The extracted viral RNA was store at −20°C till further testing.

Multiple qualitative reverse transcription real-time PCR kits for identification of SARS-CoV2 were used. These kits were already in use country wide for detection of SARS-CoV2 in human. These were (Kit 1) Real-Time Fluorescent RT-PCR Kit for detecting 2019-nCoV by BGI China (IVD &CE marked; Catalogue No. MFG030010), takes ORF 1ab gene as the target domain, (Kit 2) qRT-PCR for Novel Coronavirus (2019-nCoV) Nucleic Acid Diagnostic Kit (PCR-Flourescence Probing, IVD marked) by Sansure Biotech (Sansure Biotech Inc China, Ref No. S3102E). The kit utilizes novel coronavirus (2019-nCoV) ORF-1 gene and a conserved coding nucleocapsid protein N-gene as the target regions and finally (Kit 3) detection Kit for 2019 Novel Coronavirus RNA (PCR-Fluorescence Probing) targeting the ORF 1ab and N gene of SARS-CoV-2/2019-nCoV by Da An Gene Co., China (IVD and CE marked; Catalogue No. DA-930). Thermal cycling and results interpretation were performed as per manufacturer’s instruction.

For further confirmation these samples were also tested for envelop protein (E) gene detection using the primers / probe sets that was published by Corman V. M. et. al. (Corman et al. 2020). A 25µl reaction contain 5 µl RNA, 12.5 µl 2x reaction buffer provided with the Superscript III one step RT-PCR with platinum Taq Polymerase (Invitrogen, Darmstadt, Germany; containing 0.4 mM of each deoxyribose triphosphates (dNTP) and 3.2 mM magnesium sulphate), 1 µl of reverse transcriptase / Taq mixture from Kit and different concentration of primers and probes (Table 1). Thermal cycling was carried out at 55 C for 10 min, followed by 95 C for 3min. Then 45 cycles of 95 C for 15 sec, 58 C for 30 sec on ABI 7500 real time system (Applied Bio Systems, US. Cat # *4351104)*.

**Table 1:**
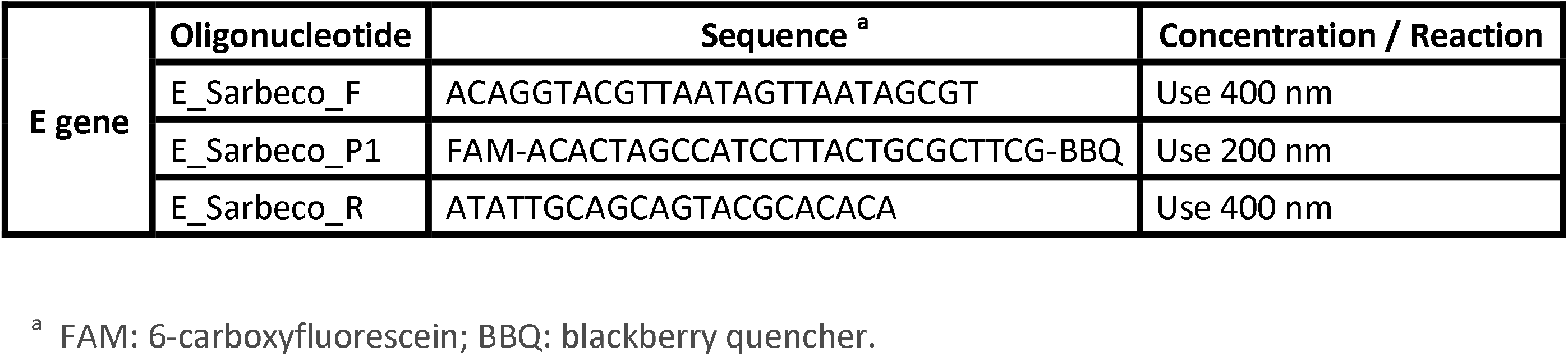
Primers and probes, real time-RT-PCR for 2019 SARS-CoV2

Genetic sequencing was based on conventional amplification of genomic RNA by using Qiagen One step RT PCR kit as described by Shirato K et al (Shirato et al. 2020). The ORF 1a gene was amplified, using a pair of primers NIID_WH-1_F501 (TTCGGATGCTCGAACTGCACC) and NIID_WH-1_R913 (CTTTACCAGCACGTGCTAGAAGG). After 1st PCR, nested PCR was performed using 2nd PCR primers (Sense: NIID_WH-1_F509(CTCGAACTGCACCTCATGG), Antisense: NIID_WH-1_R854 CAGAAGTTGTTATCGACATAGC) and 1μl of 1st PCR product under the same condition. PCR amplicons from 2nd round were purified by Qiaquick PCR purification Kit (Qiagen, Germany) and directly sequenced by using Sequencing primers (Sense: NIID_WH-1_Seq_F519 ACCTCATGGTCATGTTATGG, Antisense: NIID_WH-1_Seq_R840 GACATAGCGAGTGTATGCC) on ABI 3100 genetic analyzer using Big Dye Terminator kit V.3.0.cycle sequencing kit (ABI Foster City Canada, USA). The nucleotide sequences were assembled, edited and analyzed by Sequencher software v.4.9 (GeneCodes Incorporation, USA). The nucleotide sequence obtained was blasted against the available NCBI databank (https://blast.ncbi.nlm.nih.gov/Blast.cgi).

## Results

A total of 78 wastewater samples were collected between 12 to 18 epidemiological weeks from 38 districts across country and were tested for SARs-CoV-2 RNA. 21 wastewater samples (27%) from 13 districts were positive on RT-qPCR (Table 2).

**Table 2:** Details wastewater samples tested for SARS-CoV2 at Virology Department, National Institute of Health, Islamabad, Pakistan

| Lab# | Collection Site | Drainage Type | District | Date Collection | EPI Week | COVID-19 Results |
| --- | --- | --- | --- | --- | --- | --- |
| 186 | AQILPUR & ASLAM TOWN | PUMPING STATION | RAJANPUR | 20-Mar-20 | Week12 | NOT DETECTED |
| 189 | SUR PUL | OPEN DRAIN | QUETTA | 20-Mar-20 | Week12 | DETECTED |
| 197 | FAQIRABAD | OPEN DRAIN | KOHAT | 30-Mar-20 | Week14 | NOT DETECTED |
| 198 | HAZARA COLONY | OPEN DRAIN | KURRAM | 30-Mar-20 | Week14 | NOT DETECTED |
| 199 | COMPOSITE BUS STAND & MODC | OPEN DRAIN | DIKHAN | 30-Mar-20 | Week14 | NOT DETECTED |
| 200 | COMPOSITE SHERPAO & ZAFARABAD | OPEN DRAIN | DIKHAN | 30-Mar-20 | Week14 | NOT DETECTED |
| 201 | BAGO ROAD & BAQIRABAD | PUMPING STATION | KAMBAR | 31-Mar-20 | Week14 | NOT DETECTED |
| 202 | QILA SHEIKHUPURA & PS BAHRIAN WALA | PUMPING STATION | SHEIKHUPURA | 2-Apr-20 | Week14 | NOT DETECTED |
| 203 | RAILWAY PUL | OPEN DRAIN | QUETTA | 1-Apr-20 | Week14 | DETECTED |
| 207 | NALA BHAIR | OPEN DRAIN | SIALKOT | 2-Apr-20 | Week14 | NOT DETECTED |
| 204 | KALAPUL MURRE ROAD | OPEN DRAIN | ABOTABAD | 3-Apr-20 | Week14 | NOT DETECTED |
| 205 | HINJAL & NOORABAD | OPEN DRAIN | BANNU | 3-Apr-20 | Week14 | NOT DETECTED |
| 206 | PUMPING S.36 AHMAD NAGAR | PUMPING STATION | FAISALABAD | 3-Apr-20 | Week14 | DETECTED |
| 208 | TURWA | OPEN DRAIN | PISHIN | 4-Apr-20 | Week14 | NOT DETECTED |
| 209 | ARMY KAZIBA | OPEN DRAIN | KABDULAH | 4-Apr-20 | Week14 | NOT DETECTED |
| 210 | GULSHAN RAVI STATION | PUMPING STATION | LAHORE | 6-Apr-20 | Week15 | NOT DETECTED |
| 211 | SABZI MANDI | OPEN DRAIN | DG KHAN | 6-Apr-20 | Week15 | NOT DETECTED |
| 212 | SANGOT & THOTHAL | OPEN DRAIN | MIRPUR | 6-Apr-20 | Week15 | NOT DETECTED |
| 219 | WALKWAY NEELUM & OLD NEELUM BRIDGE | OPEN DRAIN | MUZAFARABAD | 6-Apr-20 | Week15 | NOT DETECTED |
| 213 | MIANI PUMPING STATION | PUMPING STATION | SUKKUR | 7-Apr-20 | Week15 | NOT DETECTED |
| 214 | SURAJ MIANI | PUMPING STATION | MULTAN | 8-Apr-20 | Week15 | NOT DETECTED |
| 215 | FRONTIER COLONY | OPEN DRAIN | KARACHI | 8-Apr-20 | Week15 | DETECTED |
| 216 | ORANGI NALLA | OPEN DRAIN | KARACHI | 8-Apr-20 | Week15 | NOT DETECTED |
| 217 | QASBA COLONY | OPEN DRAIN | KARACHI | 8-Apr-20 | Week15 | DETECTED |
| 218 | BANGALI PARA | OPEN DRAIN | KARACHI | 8-Apr-20 | Week15 | NOT DETECTED |
| 221 | MUHAMMAD KHAN COLONY | OPEN DRAIN | KARACHI | 9-Apr-20 | Week15 | NOT DETECTED |
| 222 | KORANGI NALLA | OPEN DRAIN | KARACHI | 9-Apr-20 | Week15 | NOT DETECTED |
| ICT-01 | SECTOR I-10/4 | OPEN DRAIN | Islamabad | 9-Apr-20 | Week15 | DETECTED |
| ICT-02 | SECTOR I-10/1 | OPEN DRAIN | Islamabad | 9-Apr-20 | Week15 | DETECTED |
| 224 | HIJRAT COLONY PIDC COLONY | OPEN DRAIN | KARACHI | 10-Apr-20 | Week15 | NOT DETECTED |
| 220 | KONRA CHINA & SPAISHTA | OPEN DRAIN | WAZIR-S | 10-Apr-20 | Week15 | NOT DETECTED |
| 223 | SABZI MANDI | OPEN DRAIN | Islamabad | 13-Apr-20 | Week16 | NOT DETECTED |
| 225 | DHOKE DALLAL | OPEN DRAIN | RAWALPINDI | 13-Apr-20 | Week16 | DETECTED |
| 226 | SAFDAR ABAD | OPEN DRAIN | RAWALPINDI | 13-Apr-20 | Week16 | DETECTED |
| 227 | MAIN DISPOSAL | PUMPING STATION | DG KHAN | 13-Apr-20 | Week16 | NOT DETECTED |
| 228 | OUTFALL STATION-G | PUMPING STATION | LAHORE | 13-Apr-20 | Week16 | DETECTED |
| 229 | OUTFALL STATION-H | PUMPING STATION | LAHORE | 13-Apr-20 | Week16 | NOT DETECTED |
| 230 | OUTFALL STATION-F | PUMPING STATION | LAHORE | 13-Apr-20 | Week16 | NOT DETECTED |
| 231 | COMPOSITE SHERPAO & ZAFARABAD | OPEN DRAIN | DIKHAN | 13-Apr-20 | Week16 | NOT DETECTED |
| 232 | COMPSITE BUS STAND & MODC | OPEN DRAIN | DIKHAN | 13-Apr-20 | Week16 | NOT DETECTED |
| 233 | RASHID MINHAS RD LAY | OPEN DRAIN | KARACHI | 13-Apr-20 | Week16 | NOT DETECTED |
| 234 | HAJI MUREED GOTH | OPEN DRAIN | KARACHI | 13-Apr-20 | Week16 | NOT DETECTED |
| 235 | TAWOOS ABAD | OPEN DRAIN | QUETTA | 12-Apr-20 | Week16 | NOT DETECTED |
| 236 | TULSIDAS PUMPING STATION | PUMPING STATION | HYDERABAD | 13-Apr-20 | Week16 | NOT DETECTED |
| 237 | KHAMISO GOTH | OPEN DRAIN | KARACHI | 14-Apr-20 | Week16 | NOT DETECTED |
| 238 | SOHRAB GOTH | OPEN DRAIN | KARACHI | 14-Apr-20 | Week16 | NOT DETECTED |
| 240 | MAKKA PUMPING STATION | PUMPING STATION | SUKKUR | 14-Apr-20 | Week16 | NOT DETECTED |
| 241 | MACHAR COLONY | OPEN DRAIN | KARACHI | 14-Apr-20 | Week16 | NOT DETECTED |
| 242 | BAGO ROAD & BAQIRABAD | PUMPING STATION | KAMBAR | 14-Apr-20 | Week16 | NOT DETECTED |
| 243 | SADDAR PUMPING STATION | PUMPING STATION | JACOBABAD | 14-Apr-20 | Week16 | NOT DETECTED |
| 244 | MASAN MULLAH | PUMPING STATION | DADU | 14-Apr-20 | Week16 | DETECTED |
| 245 | LANDHI BAKHTAWAR VILLAGE | OPEN DRAIN | KARACHI | 15-Apr-20 | Week16 | DETECTED |
| 248 | HADI PACKET | OPEN DRAIN | KILLA ABDULLAH | 15-Apr-20 | Week16 | DETECTED |
| 246 | CHAKORA NULLA | OPEN DRAIN | KARACHI | 15-Apr-20 | Week16 | NOT DETECTED |
| 239 | RAJKOT & SANSI ROAD & PEOPLE COLONY | OPEN DRAIN | GUJARANWALA | 15-Apr-20 | Week16 | NOT DETECTED |
| 252 | GANJ MOHALLA | OPEN DRAIN | ZHOB | 16-Apr-20 | Week16 | NOT DETECTED |
| 247 | CHAK & PAR HOTI | OPEN DRAIN | MARDAN | 17-Apr-20 | Week16 | NOT DETECTED |
| ICT-04 | RAWALPINDI INSTITUTE OF UROLOGY | OPEN DRAIN | RAWALPINDI | 17-Apr-20 | Week16 | DETECTED |
| ICT-06 | DHOKE KASHMIRIAN | OPEN DRAIN | RAWALPINDI | 17-Apr-20 | Week16 | DETECTED |
| 249 | PUMPING S.36 AHMAD NAGAR | PUMPING STATION | FAISALABAD | 17-Apr-20 | Week16 | NOT DETECTED |
| 250 | MILL COLONY | OPEN DRAIN | NOWSHERA | 17-Apr-20 | Week16 | DETECTED |
| 251 | SHAHEEN MUSLIM TOWN | OPEN DRAIN | PESHAWAR | 17-Apr-20 | Week16 | DETECTED |
| 253 | RASALA LINE | OPEN DRAIN | LORALAI | 17-Apr-20 | Week16 | NOT DETECTED |
| 254 | MULTAN ROAD STATION | PUMPING STATION | LAHORE | 20-Apr-20 | Week17 | DETECTED |
| 255 | AQILPUR & ASLAM TOWN | PUMPING STATION | RAJANPUR | 20-Apr-20 | Week17 | NOT DETECTED |
| 256 | SUR PUL | OPEN DRAIN | QUETTA | 20-Apr-20 | Week17 | NOT DETECTED |
| 257 | LABOUR NALA | OPEN DRAIN | DERA BUGHTI | 20-Apr-20 | Week17 | NOT DETECTED |
| 264 | KATAN PUL | OPEN DRAIN | KHUZDAR | 20-Apr-20 | Week17 | NOT DETECTED |
| 258 | WAPDA COLONY | OPEN DRAIN | NASIRABAD | 20-Apr-20 | Week17 | NOT DETECTED |
| 261 | ALI TOWN | PUMPING STATION | MULTAN | 22-Apr-20 | Week17 | NOT DETECTED |
| 262 | SILAN WALI | PUMPING STATION | SARGODAHA | 24-Apr-20 | Week17 | NOT DETECTED |
| 263 | LARA MA | OPEN DRAIN | PESHAWAR | 24-Apr-20 | Week17 | NOT DETECTED |
| 265 | KALAPUL MURRE ROAD | OPEN DRAIN | ABOTABAD | 27-Apr-20 | Week18 | DETECTED |
| 266 | FAQIRABAD | OPEN DRAIN | KOHAT | 27-Apr-20 | Week18 | DETECTED |
| 267 | SANGOT & THOTHAL | OPEN DRAIN | MIRPUR | 27-Apr-20 | Week18 | NOT DETECTED |
| 268 | LALBAGH & TIBA BAHADUR | PUMPING STATION | BAHAWALPUR | 28-Apr-20 | Week18 | NOT DETECTED |
| 269 | JATAK KILLI & TAKHTHANI | OPEN DRAIN | QUETTA | 28-Apr-20 | Week18 | DETECTED |
| 270 | KOTLA ABDUL FATAH | PUMPING STATION | MULTAN | 28-Apr-20 | Week18 | NOT DETECTED |

\* Collection Site name is designated as per drainage area or collection vicinity

Primarily, 20 wastewater samples were collected during March 20, 2020 to April 09, 2020 from 17 districts of Pakistan; 18 samples from different polio environmental sites (ES) distributed across 16 districts and 02 samples in areas with recent history of SARS-CoV2 cases from capital city Islamabad. All these samples were tested against the three commercially available SARS-CoV-2 RNA detection diagnostic kits (mentioned under materials and methods). Negative control supplied with the diagnostic kits containing the internal control was added in each sample during extraction to ensure none of the wastewater extracted RNA samples had RT-qPCR inhibition. A total of 6 samples (30 %) were positive for SARs-CoV-2 RNA, out of these all were detected on Kit 1, 2 (either one gene positive or both, results interpreted as per manufacturer’s instruction) and E gene method whereas only 4 were positive on Kit 3 (Table 2). 4 positive samples had *Cq* values between 32 to 38, whereas remaining 2 samples collected in areas with history of SARs-CoV-2 cases from Islamabad were positive on all three diagnostic kits and E gene method having *Cq* between 36 to 38 (Table 3).

**Table 3:** Wastewater samples collected during March 20, 2020 to April 09, 2020. A comparative analysis

| Sample ID | Drainage Type | Epi Week | District | Date Collection | Kit 1 (ORF 1+ Ngene) | Kit 2 (ORF 1ab+ Ngene) | Kit 3 (ORF1ab ) | Corman V. M. et. al Method (E Gene) | Final Results |
| --- | --- | --- | --- | --- | --- | --- | --- | --- | --- |
| 186 | PUMPING STATION | Week 12 | RAJANPUR | 20-Mar-20 | ND | ND | ND | ND | ND |
| 189 | OPEN DRAINAGE | Week 12 | QUETTA | 20-Mar-20 | + | + | ND | + | DETECTED |
| 197 | OPEN DRAINAGE | Week 14 | KOHAT | 30-Mar-20 | ND | ND | ND | ND | ND |
| 198 | OPEN DRAINAGE | Week 14 | KURRAM | 30-Mar-20 | ND | ND | ND | ND | ND |
| 199 | PUMPING STATION | Week 14 | DIKHAN | 30-Mar-20 | ND | ND | ND | ND | ND |
| 200 | OPEN DRAINAGE | Week 14 | DIKHAN | 30-Mar-20 | ND | ND | ND | ND | ND |
| 201 | OPEN DRAINAGE | Week 14 | KAMBAR | 31-Mar-20 | ND | ND | ND | ND | ND |
| 202 | PUMPING STATION | Week 14 | SHEIKHUPURA | 1-Apr-20 | ND | ND | ND | ND | ND |
| 203 | OPEN DRAINAGE | Week 14 | QUETTA | 1-Apr-20 | + | + | + | + | DETECTED |
| 204 | OPEN DRAINAGE | Week 14 | ABOTABAD | 3-Apr-20 | ND | ND | ND | ND | ND |
| 205 | OPEN DRAINAGE | Week 14 | BANNU | 3-Apr-20 | + | + | + | + | DETECTED |
| 206 | PUMPING STATION | Week 14 | FAISALABAD | 3-Apr-20 | + | + | ND | + | DETECTED |
| 207 | OPEN DRAINAGE | Week 14 | SIALKOT | 2-Apr-20 | ND | ND | ND | ND | ND |
| 208 | OPEN DRAINAGE | Week 14 | PISHIN | 2-Apr-20 | ND | ND | ND | ND | ND |
| 209 | OPEN DRAINAGE | Week 14 | KABDULAH | 4-Apr-20 | ND | ND | ND | ND | ND |
| 210 | PUMPING STATION | Week 15 | LAHORE | 6-Apr-20 | ND | ND | ND | ND | ND |
| 211 | PUMPING STATION | Week 15 | DGKHAN | 6-Apr-20 | ND | ND | ND | ND | ND |
| 212 | OPEN DRAINAGE | Week 15 | MIRPUR | 6-Apr-20 | ND | ND | ND | ND | ND |
| ICT-01 | OPEN DRAINAGE | Week 15 | ISLAMABAD | 9-Apr-20 | + | + | + | + | DETECTED |
| ICT-02 | OPEN DRAINAGE | Week 15 | ISLAMABAD | 9-Apr-20 | + | + | + | + | DETECTED |

\* Collection Site name is designated as per the drainage area or collection vicinity ND Not detected

A centrifugation step was also introduced before Viral RNA extraction to increase viral RNA yield. A decrease in *Cq* value has been observed, describing an increase in viral RNA concentration (Table 4). Furthermore, 56 samples collected and received in laboratory from April 6, 2020 to April 28,2020 for polio diagnostics were also tested for detection of SARs-CoV-2, out which 14 (24%) were positive. One sample was collected from the drain of a Rawalpindi Institute of Urology (RIU), a COVID-19 patient quarantine center, while other from a drain having catchment area with recent history of COVID-19 patients. Both collection sites are in district Rawalpindi. These samples were found positive for SARs-COV-2 RNA on RT-qPCR.

**Table 4:**
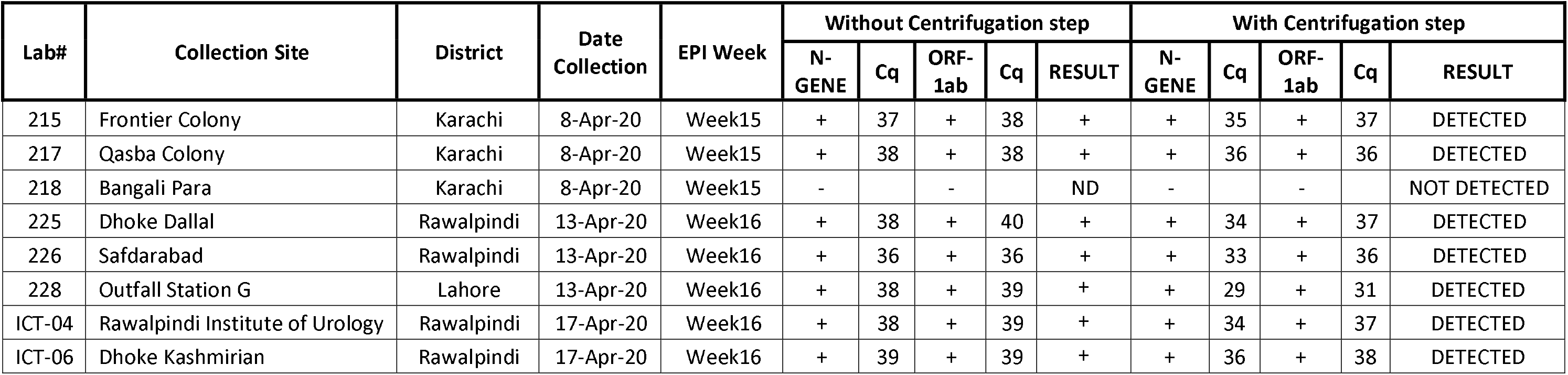
Sample preparation without and with centrifugation before viral RNA Extraction. Comparison among the *Cq* values

The sample collected from the drain of RIU, Rawalpindi was also subjected for partial sequencing of SARs-CoV-2 ORF-1a. Nucleotide sequence of partial SARs-CoV-2 ORF 1a is submitted in gene bank under accession number MT539157.

## Discussion

The role of environmental surveillance in supporting Global Polio Eradication Initiative has already been acknowledged (Kroiss et al. 2018). The environmental surveillance can be used as supplemental tool for detection pathogens circulating within the community. Wastewater provides a near-real-time data as it constantly collects feces, urine and traces of sputum that can contain SARs-CoV-2 shed by the infected individuals. Viral load estimation in COVID-19 positive patients are still uncertain, however, recently N. Zhang *et al*. suggests levels as high as 600,000 viral genomes per ml of fecal material (Wu FQ 2020). Similarly another study reported approximately 30,00,000 viral particles in a single fecal sample (Wu FQ 2020). A previously published study on coronavirus reported that it remained infectious in water and sewage for days to weeks. Researchers reported time required for 99% reduction of virus infectivity was several days at room temperature in pure water or wastewater (Guangbo Qu 2020). This adds another potential supplemental detection source of SARs-CoV-2 in communities.

In this study, we investigated the presence of SARs-CoV-2 RNA in wastewater using the existing poliovirus environment surveillance network that can be used in future as an early warning system for the dependent area. This definitely needs further evaluation and discussions; however, this seems to have a very interesting utilization in epidemiology. A total of 21 out of 78 positive wastewater samples for SARs-CoV-2 RNA clearly indicates viral RNA shedding in stool of infected individuals. Currently, there is no evidence of infection transmission of SARs-COV-2 or related SARs-Corona via wastewater (Ahmed et al. 2020).

We used three commercially available kits and Published E-Gene detection assay for surety and confirmatory of positivity. Analyzing table 3, samples collected from Quetta district at two different time intervals indicates COVID-19 prevalence and surge in infected individuals. This can be assumed from the decrease in *Cq* value in wastewater sample collected two weeks after. Detection of SARs-COV-2 RNA in two specific wastewater samples collected from areas with recent history of COVID-19 patients clearly explain that wastewater testing for COVID-19 can be used as an early warning system. Likewise, SARs-COV-2 RNA positive samples from RIU, Rawalpindi and an area in Rawalpindi with COVID patients further strengthen the findings and use of this tool in future. This surveillance system can picks up vast majority of infected individuals with SARs-CoV-2 who do not present symptoms for the disease (Ahmed et al. 2020). Furthermore, sequence data of partial ORF 1a generated from ICT-04 also reinforce our findings that SARs-CoV-2 can be detected in wastewater in Pakistan. Interestingly, the additional extra centrifugation step before viral RNA extraction seems to be encouraging in increasing the yield of viral RNA. This can be obvious from the data presented in table 4.

The surveillance through wastewater can be useful in remote or confined communities, however, further studies are needed on virus concentration and detection assay to increase the sensitivity. This has an epidemiologic potential for early detection of high burden area in advance; and heavily populated areas where door-to-door tracing may not be possible. This may also be more relevant to the developing countries with limited molecular testing. Development of highly sensitive assay will be an indicator for virus monitoring and to provide early warning signs.

## Conclusion

SARS-CoV-2 detection in wastewater using RT-qPCR assay, confirmed by sequencing, is a milestone in the field of epidemiology. The study finding indicates that environmental surveillance through wastewater could be used as early warning system to monitor viral tracking and circulation in cities with lower COVID-19 disease burden or setting where person to person testing is limited. The virus concentration and detection method in wastewater needs attention to increase sensitivity of detection of SARs-CoV-2 in wastewater.

## Data Availability

All Data used in the present study included in the manuscript.

---

**Figure 1:**
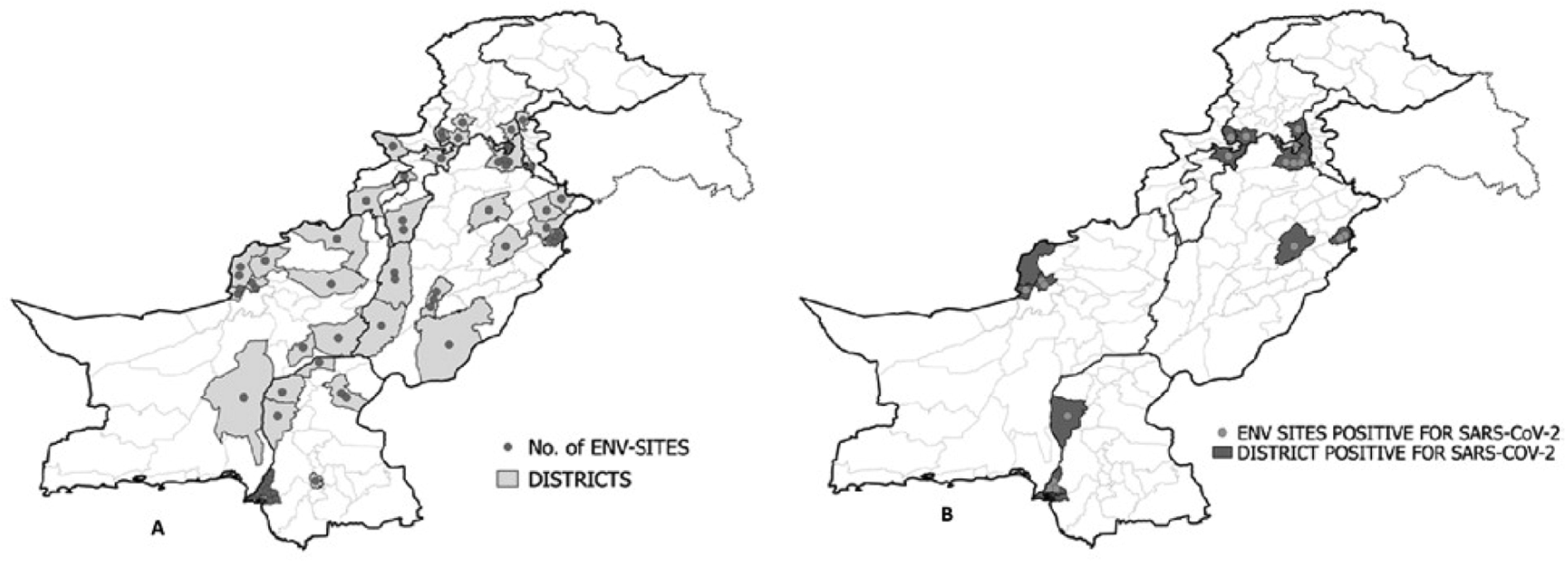
**A**. Map indicating environmental sampling sites. Each purple dot represents a wastewater collection site **B**. Map indicating the red marked districts with SARs-CoV-2 positive wastewater samples. Each orange dot represents a positive wastewater sample

**Figure 2:**
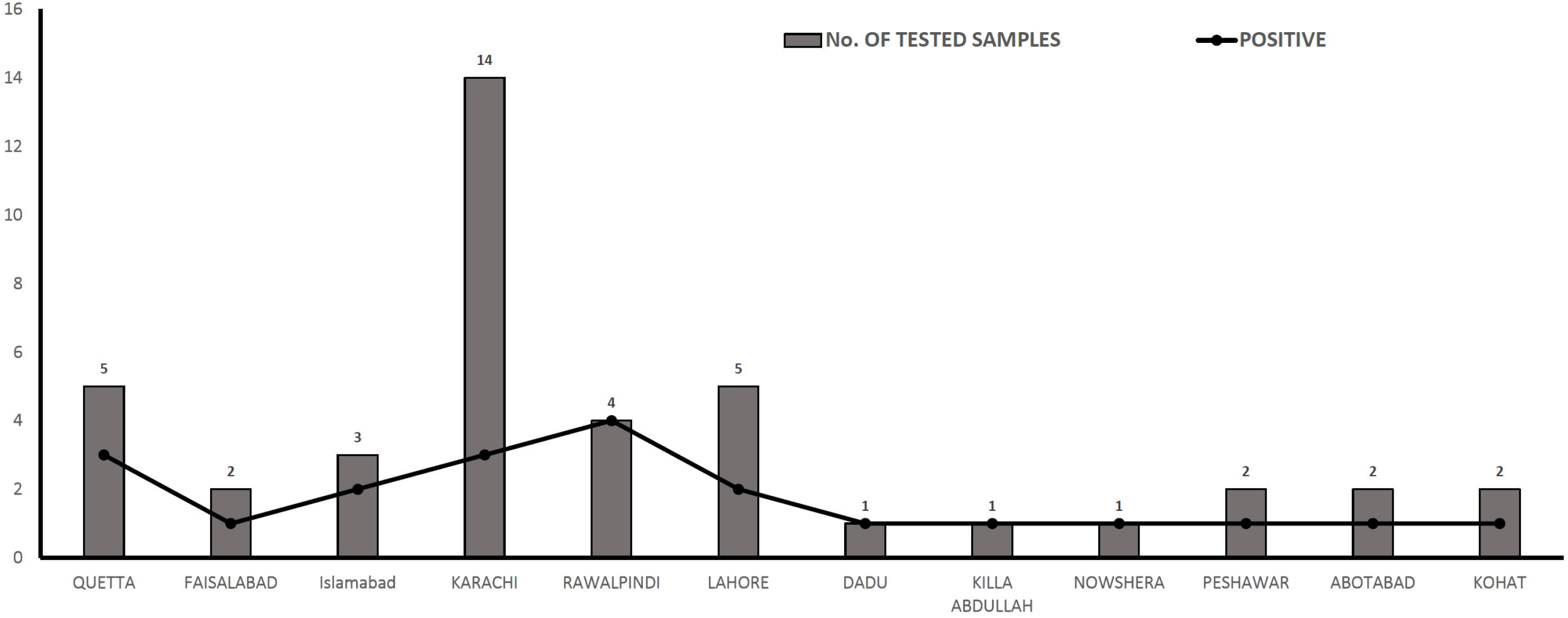
Graphical representation of positive samples among tested samples.

